# A Decade’s Experience Performing Forensic Medical and Psychologic Evaluations for Pediatric Asylum Seekers in the United States

**DOI:** 10.1101/2023.09.10.23295337

**Authors:** Marina Plesons, Haley Hullfish, Priyashma Joshi, Stephen Symes, Anjali Saxena

## Abstract

**Background:** The number of children seeking asylum in the US has increased substantially in recent years. To add to the limited evidence base related to children seeking asylum in the US, this paper describes the sociodemographic characteristics, nature of human rights violations, and guardianship status of the children served by the Human Rights Clinic of Miami from 2010-2021.

**Methods:** A retrospective study was conducted on affidavits for pediatric patients seeking forensic medical and psychological evaluation at the Human Rights Clinic of Miami from 2010 to 2021. We determined trends among sociodemographic characteristics and the nature of human rights violations among these patients and used logistic regression to determine factors associated with having a guardian present at the evaluation.

**Results:** Of the total 64 pediatric patients who received a forensic medical and psychological evaluation, over half were male (56%) and two-thirds were aged 15-17. Honduras was the most common country of origin (53%). Physical violence was the most reported human rights abuse (72%) followed by gang violence (53%), sexual violence (22%), and political violence (3%). The majority of children (87%) reported being detained at the border upon entry into the USA. Only 31% of pediatric patients had a guardian present during the evaluation, with guardianship significantly less likely for older patients (p <0.05).

**Conclusions:** These findings identified specific challenges unique to the pediatric population, including lack of guardianship among older children. Future efforts may include the development of formal pediatric-specific guidelines for conducting evaluations of children seeking asylum.

## Introduction

In 2022, over 300,000 children were arrested by border patrol at the United States (US)-Mexico border, representing a five-fold increase since 2008 [1]. Furthermore, there was a seventeen-fold increase in the number of unaccompanied children encountered by border control officers between 2008 and 2019 [2].

Many of these individuals come to the US to seek asylum due to persecution or fear of persecution due to their race, gender, religion, nationality, membership in a particular social group, and/or political opinion. Although the odds of being granted asylum are strongly affected by the quality of their legal representation and by the immigration judge to which they are assigned, the US government does not provide an attorney for individuals seeking asylum [3, 4]. For example, the percent of applications that were granted asylum in Miami from 2017-2022 ranged from 0.9% to 27.9% based on which judge presided over the case [5]. The number of pending immigration cases has also increased six-fold in the past ten years, totaling nearly 2 million in 2022, with an average wait time of 795 days [6].

Forensic medical and psychological evaluations by health professionals that corroborate claims of persecution increase an individual’s likelihood of being granted asylum [7-13]. During these evaluations, health professionals assess the individual’s overall health and identify signs and symptoms of human rights abuse, and then summarize this evidence in an affidavit to be used in court in the individual’s application for asylum. In 1994, the Office of the High Commissioner for Human Rights published the *Istanbul Protocol: Manual on the Effective Investigation and Documentation of Torture and Other Cruel, Inhuman or Degrading Treatment or Punishment*, later updated in 2004 and 2022, to guide health professionals on conducting such evaluations [14]. Various manuals, guidelines, and toolkits have been established since then to provide additional support to health professionals [15-19].

Most of these resources focus on adults but recent preliminary efforts have been made to develop specific guidance on conducting forensic evaluations for children [20, 21]. However, there is a need for more involvement from the limited medical community involved in this work, as there is lack of consensus on issues such as the impact of trauma on pediatric development and best practices for minimizing re-traumatization of children during the evaluation process [22]. Likewise, a national survey conducted in 2020 identified only 28 providers who reported performing humanitarian relief evaluations for children seeking asylum in the USA, pointing to the pressing need to train more health professionals in the forensic evaluation of children [22].

The Human Rights Clinic of Miami is an independent collection of medical students, residents, and attending physicians who provide impartial medical and psychological evaluations for asylum seekers in the US. As of November 2022, Miami-Dade County had 105,316 residents with pending immigration court deportation cases, the most of any US city [23]. The percentage of applications granted asylum in Miami is consistently below the national average; it decreased from a high of 41.5% in 2012 to 8.5% in 2020 and increased again to 24.8% in 2022 (Fig 1) [24].

**Figure 1.**
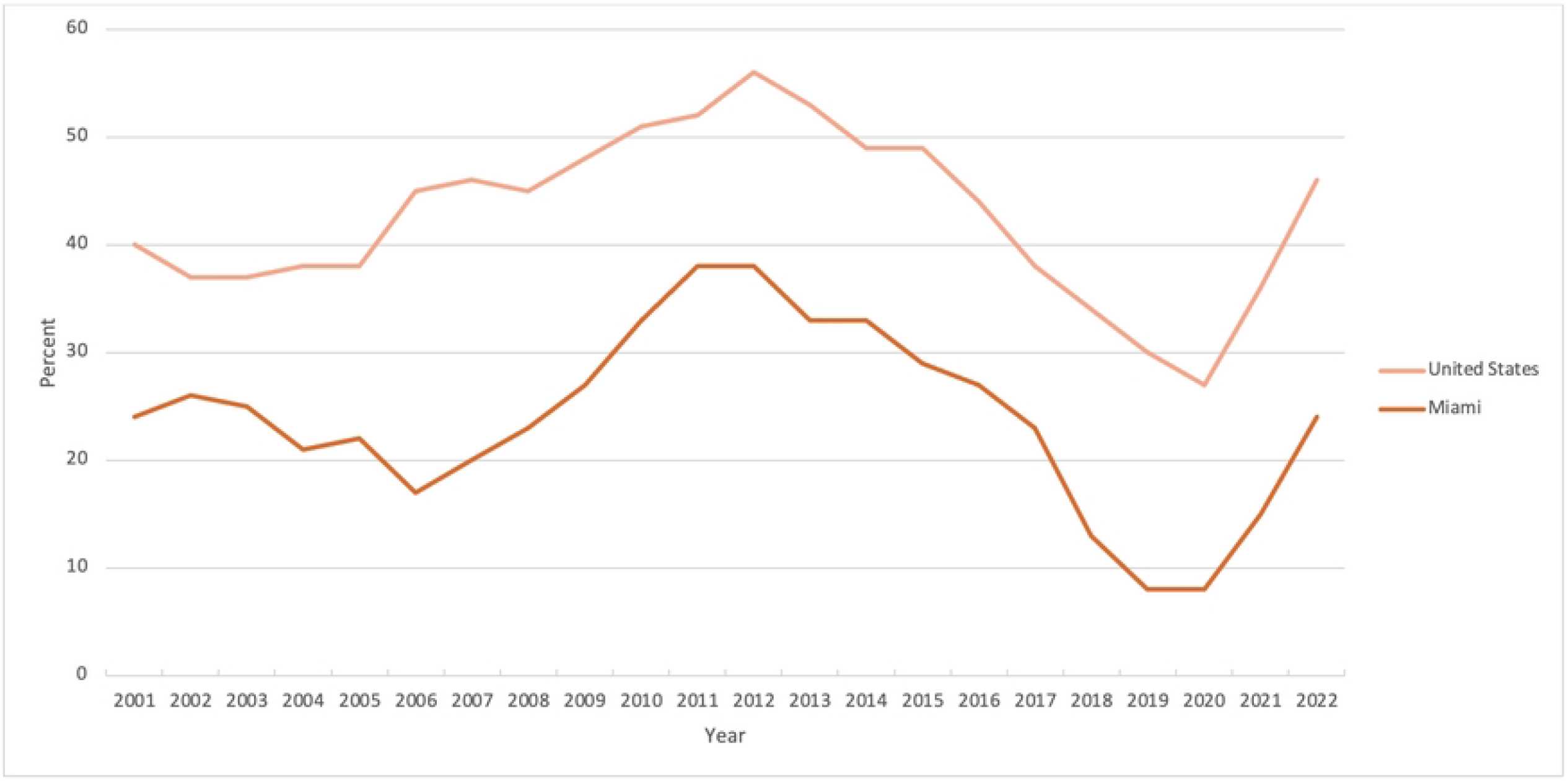
Percentage of applicants granted asylum, by fiscal year.

This paper seeks to add to the limited evidence base regarding forensic medical and psychological evaluations of children seeking asylum by describing the sociodemographic attributes and guardianship status of the children served by the Human Rights Clinic of Miami from 2010-2021 so that physicians are better able to assess this population in the future.

## Methods

A retrospective chart review was conducted of all affidavits prepared by the Human Rights Clinic of Miami for pediatric patients from 2010 – 2021. Consent for use of their information in research activities conducted by the Human Rights Clinic of Miami was sought from participants’ parents/guardians at the time of their evaluation, as was assent from the participants themselves. Before 2020, this consisted of written consent. In 2020 and 2021, this consisted of oral consent as evaluations were conducted via Zoom due to restrictions related to the COVID-19 pandemic; oral consent was witnessed and documented by at least two other staff members of the Human Rights Clinic of Miami. Data was accessed in January and February 2022 and was organized using REDCap electronic data capture tools hosted at the University of Miami. During data collection, the authors had access to information that could identify individual participants; however, efforts were made to ensure the privacy of this information and this information was not extracted from the source. Statistical analyses were performed using SPSS (IBM Corp. Released 2021). Descriptive statistics were used to describe the sociodemographic characteristics and nature of human rights violations of the children who received services from the Human Rights Clinic of Miami, and logistic regression was used to identify factors associated with the likelihood of children having a guardian present during the medical and psychological evaluation.

## Results

From 2010-2021, the Human Rights Clinic of Miami conducted 64 forensic medical and psychological evaluations for pediatric patients, constituting 17% of the evaluations conducted by the Clinic during that time (Table 1). Just over half of the children were male (56%). Two-thirds of the children were 15-17 years old, one-quarter were 11-14 years old, and 6% were 5-10 years old; no children were less than 5 years old. Honduras was the most common country of origin (53%), followed by Guatemala (28%) and El Salvador (16%). The majority of pediatric patients reported achieving some primary (elementary) school in their country of origin (63%), while 29% reported achieving some secondary (high) school, and 8% reported receiving no education.

**Table 1.** Demographics of the Pediatric Patients at the Human Rights Clinic of Miami.

| Characteristic | N |  | Frequency (n) | Percentage (%) |
| --- | --- | --- | --- | --- |
| Gender | 64 | Female | 28 | 43.8 |
|  |  | Male | 36 | 56.3 |
| Age (years) | 64 | <5 | 0 | 0 |
|  |  | 5-10 | 4 | 6.3 |
|  |  | 11-14 | 17 | 26.5 |
|  |  | 15-17 | 43 | 67.2 |
| Race | 6 | Black | 2 | 33.3 |
|  |  | Indigenous | 4 | 66.6 |
| Ethnicity | 62 | Hispanic | 62 | 100 |
| Country of origin |  | El Salvador | 10 | 15.6 |
|  |  | Guatemala | 18 | 28.1 |
|  |  | Haiti | 1 | 1.6 |
|  |  | Honduras | 34 | 53.1 |
|  |  | United States of America | 1 | 1.6 |
| Highest level of education in home country | 52 | None | 4 | 7.7 |
|  |  | Some primary (elementary) school | 33 | 63.5 |
|  |  | Some secondary (high) school | 15 | 28.8 |
| Type of human rights abuse | 64 | Gang | 34 | 53.1 |
|  |  | Political | 2 | 3.1 |
|  |  | Physical violence | 46 | 71.9 |
|  |  | Sexual violence | 14 | 21.9 |
|  |  | Other | 1 | 1.6 |
| Detained at the border | 37 | Yes | 32 | 86.5 |
|  |  | No | 5 | 13.5 |
| Number of people in house | 50 | <5 | 32 | 64.0 |
|  |  | 5-10 | 17 | 34.0 |
|  |  | >10 | 1 | 2.0 |
| Relations with people in home | 52 | Family | 50 | 96.2 |
|  |  | Mixed | 2 | 3.8 |
| Currently in school | 50 | Yes | 48 | 96.0 |
|  |  | No | 2 | 4.0 |
| Speaks English | 51 | Yes | 14 | 27.5 |
|  |  | No | 37 | 72.5 |
| Guardian present during medical and psychological evaluation | 58 | Yes | 18 | 31.0 |
|  |  | No | 40 | 69.0 |
| Guardian relationship | 18 | Mother | 15 | 83.3 |
|  |  | Father | 1 | 5.6 |

Regarding the human rights abuses that the children experienced in their country of origin, physical violence was most reported (72%), followed by gang violence (53%), sexual violence (22%), and political violence (3%). When disaggregated by gender (Fig 2), more boys reported experiencing gang violence than girls, while nearly all those who reported experiencing sexual and political violence were girls. Physical violence was reported by roughly the same number of boys and girls. When disaggregated by age (Fig 3), 5-10-year-olds reported experiencing physical violence most commonly, followed by gang violence; no 5-10-year-olds reported experiencing sexual or political violence. Most of those reporting sexual violence were 15-17 years of age.

**Figure 2.**
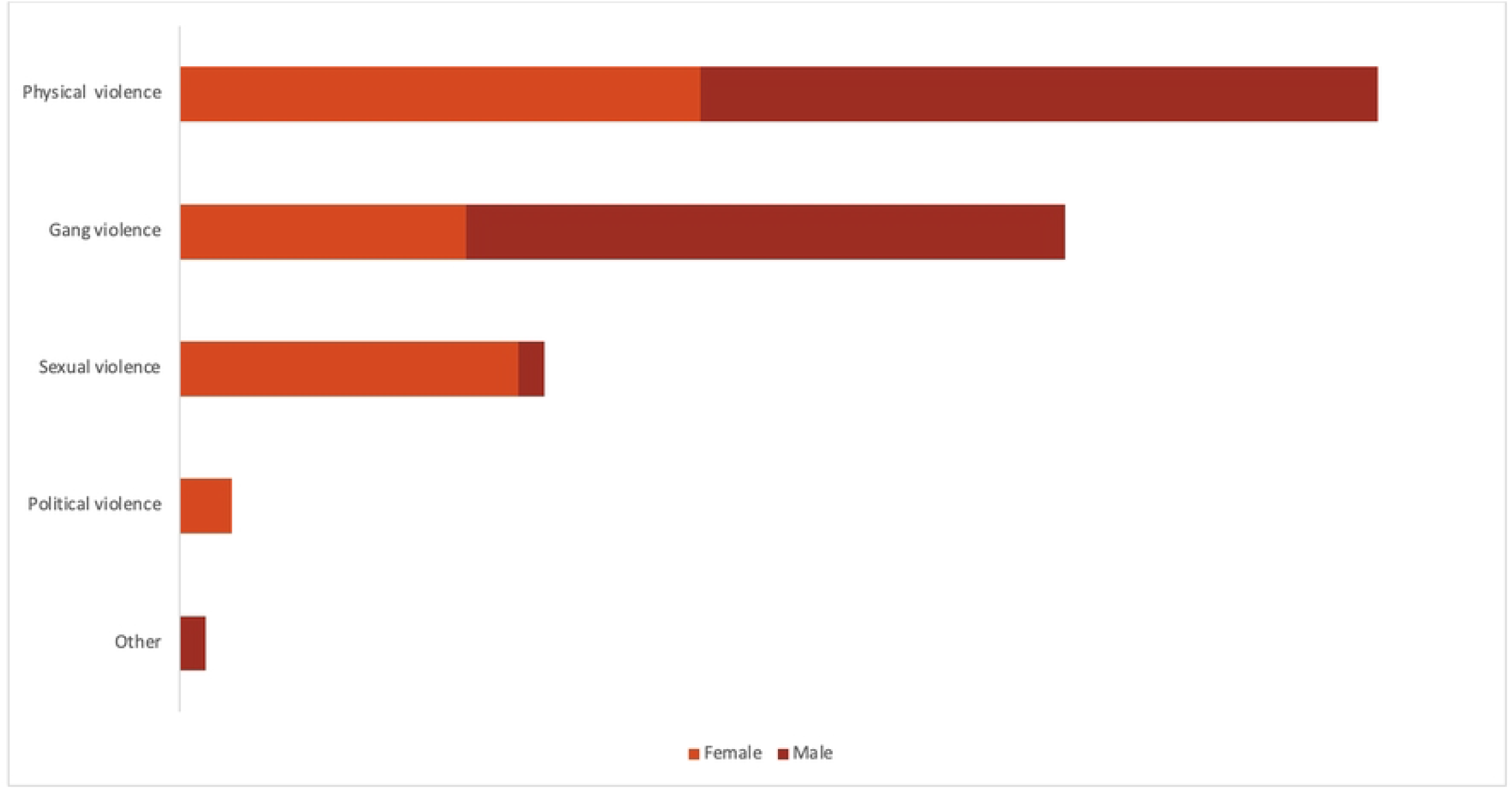
Type of human rights abuse experienced in country of origin, by gender.

**Figure 3.**
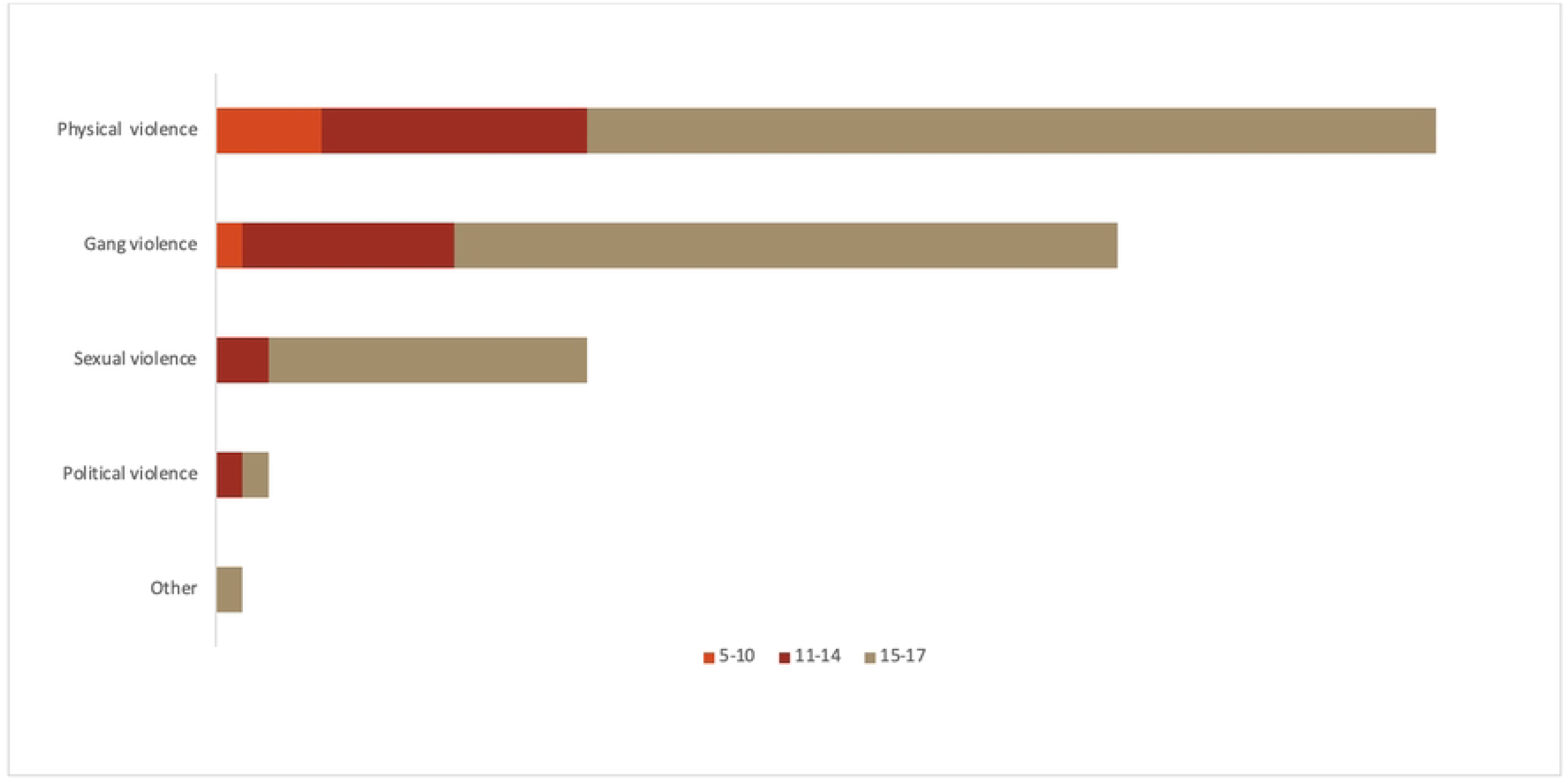
Type of human rights abuse experienced in country of origin, by age.

The majority of children (87%) reported being detained at the border upon entry into the USA. Regarding their current conditions in Miami, 64% reported living in a home with fewer than 5 people, while 34% reported living in a home with 5-10 people. Approximately all the children (96%) were currently attending school, but nearly three-quarters (73%) reported they did not speak English.

Only one in three pediatric patients (31%) had a guardian present during the medical and psychological evaluation. Of the guardians present, the majority were mothers (83%), followed by siblings (11%) and fathers (6%). When disaggregated by age (Fig 4), the percentage of children with a guardian present decreased from 100% of 5-10-year-olds, to approximately half of 11-14-year-olds, to only 20% of 15-17-year-olds. A logistic regression was performed to ascertain the effects of age, gender, and country of origin on the likelihood that a guardian was present. The model was statistically significant (χ2(6) = 16.568, p < 0.05), and explained 35.0% (Nagelkerke R2) of the variance in guardianship, correctly classifying 79.3% of cases. Increasing age was associated with a decreased likelihood of having a guardian present. Neither gender nor country of origin were significant predictors.

**Figure 4.**
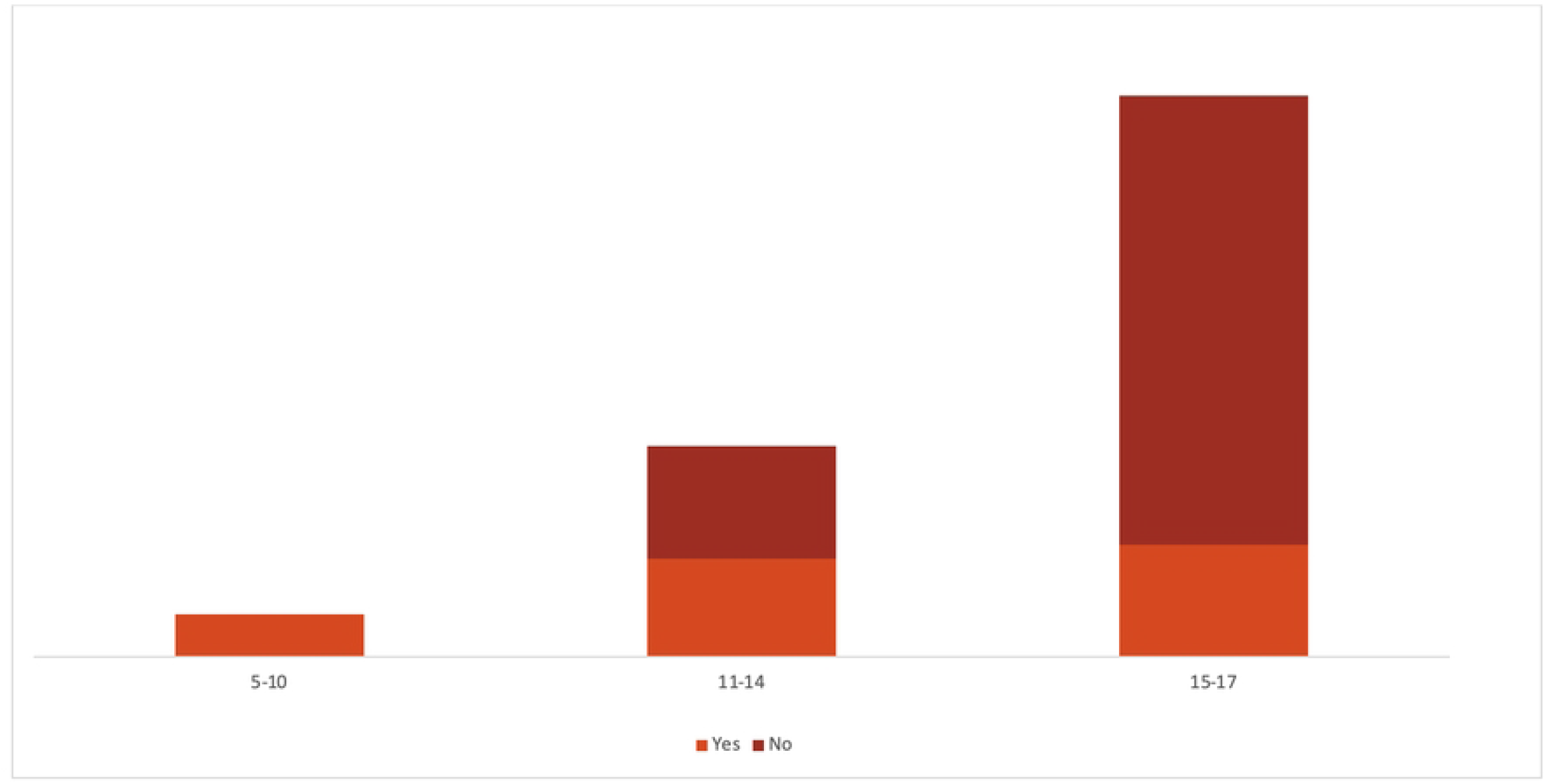
Guardian present during medical and psychological evaluation, by age.

## Discussion

In the last decade, there has been a substantial rise in unaccompanied children entering the USA [2]. Children seeking asylum present with a distinct set of physical and psychological needs compared to their adult counterparts. Unfortunately, there are still very few resources targeted towards pediatric asylum seekers [25]. Medical and psychological affidavits are strongly associated with positive outcomes in asylum cases, especially for children [13]. The Human Rights Clinic of Miami serves this population by providing such affidavits for their asylum cases, as well as medical and psychological referrals and linkage to relevant social services. Over the past ten years, pediatric patients have constituted nearly 20% of the total patients served by the Human Rights Clinic of Miami. Therefore, it is vital that we understand their unique demographics and needs to develop standardized and optimized procedures that cater specifically to this vulnerable population.

The type of violence encountered by pediatric patients in the Human Rights Clinic creates a strong argument for asylum. Our data demonstrate that most pediatric patients encountered physical violence, followed by gang, sexual, and political violence. When considering gender, both male and female pediatric patients were equally vulnerable to physical violence, but males experienced more gang violence, while females experienced more sexual and political violence. Regarding age, children aged 5–10 years were more likely to experience physical violence, while those aged 15– 17 years were more likely to experience sexual violence. This demographic data can inform health professionals on how to appropriately screen patients based on their gender and age group. Given the frequency of sexual violence in female adolescent patients for example, screenings in this population should incorporate questions on sexual violence regardless of their documented history.

This study also provides valuable insight into factors associated with guardianship of children seeking asylum. For children under 18 years old, the Human Rights Clinic of Miami always requests that a legal guardian be present for the evaluation to help to advocate on the child’s behalf, provide emotional support, protect patient autonomy, and fill in gaps in the story. Evaluations commonly involve triggering discussions of violence, abuse, and trauma, so it is crucial to have a guardian present. Furthermore, pediatric patients may not remember or be able to accurately describe relevant details about their experiences, especially those that occurred when they were very young; in such cases, a guardian can help by providing supplementary information. However, it is often difficult for guardians to attend the evaluation for various reasons, including work, childcare, and financial restraints; additionally, many children traveled to the US as unaccompanied minors and are in various stages of the process of being receiving a court-appointed guardian. Therefore, evaluations often must proceed without a guardian present, as the benefit of completing the medical and psychological evaluation in the legal time frame of their asylum case takes precedence. As demonstrated by this study, only 31% of the HRC’s pediatric patients had a guardian present at their evaluation. Older age was associated with not having a guardian present, as most 15–17-year-olds were not accompanied by a guardian. As such, guardianship demonstrates a unique problem in the evaluation of children seeking asylum that must be addressed in a more concerted manner in the future. This could include obtaining consent from a guardian before the evaluation or over the phone at the time of evaluation, obtaining assent from the child, working with the patient and lawyer further in advance of the evaluation to ensure a guardian is present, and identifying other sources of support for our patients so they can receive the care they need.

This study was conducted at a single clinic with a limited number of pediatric evaluations; as such, it presents certain limitations. Miami has a unique immigrant and asylee population, with most individuals originating from Central and South America; therefore, the results of this study may not be generalizable to the entire US. More data is necessary to place the demographic trends of pediatrics patients of the Human Rights Clinic of Miami with those served by other institutions and individuals across the country. In addition, as it is run by students at the University of Miami Miller School of Medicine, the staff of the Human Rights Clinic of Miami turn over each year; therefore, several students have inputted data into its record-keeping system over the last decade. This leads to inconsistent data input, although the data collection is standardized through REDCap.

## Conclusions

The Human Rights Clinic of Miami aims to provide medical and psychological evaluations for individuals seeking asylum in the US, while protecting their autonomy and safety. The latter goal is especially emphasized when evaluating children, a vulnerable population with unique needs. This study established the demographics of the pediatric patients seen by the Human Rights Clinic of Miami over the last ten years and identified specific challenges unique to the pediatric population. Future research may investigate whether factors such as guardianship affect the outcome of the asylum case, and future efforts may include the development of formal pediatric-specific guidelines for conducting medical and psychological evaluations of children seeking asylum.

## Data Availability

The datasets generated and/or analyzed during the current study are not publicly available due ethical considerations but are available from the corresponding author on reasonable request.

## Acknowledgements

We thank the staff members of the Human Rights Clinic of Miami for their efforts in conducting evaluations for the clinics, members of the legal team working concurrently with the HRC, and most importantly, the patients and their family members for sharing their stories.

